# A comparison of four epidemic waves of COVID-19 in Malawi; an observational cohort study

**DOI:** 10.1101/2022.02.17.22269742

**Authors:** Catherine Anscombe, Samantha Lissauer, Herbert Thole, Jamie Rylance, Dingase Dula, Mavis Menyere, Belson Kutambe, Charlotte van der Veer, Tamara Phiri, Ndaziona P. Banda, Kwazizira S. Mndolo, Kelvin Mponda, Chimota Phiri, Jane Mallewa, Mulinda Nyirenda, Grace Katha, Henry Mwandumba, Stephen B. Gordon, Kondwani C. Jambo, Jennifer Cornick, Nicholas Feasey, Kayla G. Barnes, Ben Morton, Philip M. Ashton, Blantyre COVID-19 Consortium

## Abstract

**Background:** Compared to the abundance of clinical and genomic information available on patients hospitalised with COVID-19 disease from high-income countries, there is a paucity of data from low-income countries. Our aim was to explore the relationship between viral lineage and patient outcome.

**Methods:** We enrolled a prospective observational cohort of adult patients hospitalised with PCR-confirmed COVID-19 disease between July 2020 and March 2022 from Blantyre, Malawi, covering four waves of SARS-CoV-2 infections. Clinical and diagnostic data were collected using an adapted ISARIC clinical characterization protocol for COVID-19. SARS-CoV-2 isolates were sequenced using the MinION™ in Blantyre.

**Results:** We enrolled 314 patients, good quality sequencing data was available for 55 patients. The sequencing data showed that 8 of 11 participants recruited in wave one had B.1 infections, 6/6 in wave two had Beta, 25/26 in wave three had Delta and 11/12 in wave four had Omicron. Patients infected during the Delta and Omicron waves reported fewer underlying chronic conditions and a shorter time to presentation. Significantly fewer patients required oxygen (22.7% [17/75] vs. 58.6% [140/239], p<0.001) and steroids (38.7% [29/75] vs. 70.3% [167/239], p<0.001) in the Omicron wave compared with the other waves. Multivariable logistic-regression demonstrated a trend toward increased mortality in the Delta wave (OR 4.99 [95% CI 1.0-25.0 p=0.05) compared to the first wave of infection.

**Conclusions:** Our data show that each wave of patients hospitalised with SARS-CoV-2 was infected with a distinct viral variant. The clinical data suggests that patients with severe COVID-19 disease were more likely to die during the Delta wave.

**Summary:** We used genome sequencing to identify the variants of SARS-CoV-2 causing disease in Malawi, and found that each of the four waves was caused by a distinct variant. Clinical investigation suggested that the Delta wave had the highest mortality.

## Introduction

There is limited COVID-19 genomic surveillance data from low income countries such as Malawi [1]. Genomic surveillance data supports the development of contextually relevant and effective national, regional and international public health interventions [2]. For patients with severe disease, little is known about the impact of viral variants on disease severity in these resource constrained settings where there is frequently a high prevalence of concomitant HIV-infection. Early data from South Africa suggested that the emergence of the SARS-CoV-2 omicron variant of concern (VOC) was associated with reduced disease severity [3], but there is a paucity of data from neighbouring countries in the region.

Genomic sequencing is a vital tool to inform strategies for an effective COVID-19 care and treatment response. The early release of the Wuhan-1 genome sequence [4] enabled the development of specific diagnostic tests [5] and the design of mRNA vaccines, used to great success in high-income countries [6,7]. The evolution of the virus has led to the emergence of lineages designated as VOCs, which are defined using genome sequencing and the widespread use of genomic surveillance to inform public health strategy has been a defining feature of the pandemic [8,9]. Early data on the emergence of VOCs has enabled policy makers to rapidly implement public health responses to constrain disease spread; prepare health systems (e.g. increased oxygen provision; opening more hospital beds; and increasing testing); and to select optimal vaccines and therapies [10]. In Malawi, Blantyre is the commercial hub with high detected rates of COVID-19 disease [11]. We previously deployed the WHO-accredited International Severe Acute Respiratory and Emerging Infection Consortium (ISARIC) clinical characteristation protocol at Queen Elizabeth Central Hospital (QECH) to patients admitted with suspected COVID-19 disease [12]. However, this cohort completed in September 2020; and did not include pathogen genome sequencing.

In this study we determined SARS-CoV-2 genome sequences from swabs collected from adult patients admitted to Queen Elizabeth Central Hospital (QECH) with PCR-confirmed and symptomatic COVID-19 during four sequential waves of the pandemic. Our aim was to explore the relationship between viral lineage and patient outcome in southern Malawi using an international clinical characterisation protocol. Based on emerging data from other settings [13–16], we hypothesised that there would be increased disease severity for patients with confirmed Delta disease.

## Methods

### Study design and recruitment

We prospectively recruited adult patients (>18 years old) using the tier one sampling strategy from the International Severe Acute Respiratory and Emerging Infection Consortium (ISARIC) Clinical Characterisation Protocol (CCP) [17], as previously described [12]. Patients were recruited at Queen Elizabeth Central Hospital (QECH), Blantyre, Malawi, the largest referral hospital in southern Malawi. For this study, only patients admitted to hospital with severe acute respiratory infection and a positive SARS-CoV-2 PCR test (defined as a Ct <40) were included. Patients (or personal consultee if the patient lacked capacity) with a severe acute respiratory infection (SARI) were consecutively approached for informed consent with an aim to recruit within 72 hours of hospital admission. Respiratory samples (combined nasopharyngeal and oropharyngeal swab) and peripheral venous blood samples were collected at recruitment. SARS-CoV-2 PCR diagnostic testing was carried out on the swab samples, as previously described [12]. Waves (W) of SARS-CoV-2 were defined with reference to nationally reported COVID-19 figures (W1: 04/2020 – 10/2020, W2: 11/2020 – 03/2021; W3: 04/2021 – 08/2021; and W4: 12/2021 – 03/2022). COVID-19 vaccine became available in Malawi from 10^th^ March 2021 [18].

During the recruitment period, patients with COVID-19 were treated on wards capable of providing continuous oxygen therapy, but without capacity for invasive mechanical ventilation, intensive care facilities, continuous positive airways pressure (CPAP) or high flow oxygen. All patients received protocolised standard care depending on the severity, including oxygen, steroids and antibiotics as previously described [19]. Clinical and treatment parameters were recorded using the ISARIC standardised case report form. Participants were followed up until death, discharge or transfer to another facility.

Study protocols were approved by the Malawi National Health Science Research Committee (NHSRC, 20/02/2518 and 19/08/2246) and Liverpool School of Tropical Medicine Research Ethics Committee (LSTM REC, 20/026 and 19/017). We have included a reflexivity statement detailing how equitable partnership was promoted within our collaboration in the Supplementary Material.

### SARS-CoV-2 molecular biology and genome sequencing

Samples were extracted using the Qiasymphony-DSP mini kit 200 (Qiagen, UK) with offboard lysis or manually using the Qiagen mini viral extraction kit. Samples were then tested using the CDC N1 assay to confirm the Ct values before sequencing. ARTIC protocol V2 sequencing protocol was used until June 2021, after which we switched to the V3 protocol. ARTIC version 3 primers were used for the tiling PCR until we switched to the University of Zambia (UNZA) primer set that provided better results for Delta VOC in August 2021 (data not shown) [20]. Initially two primer pools were used, however a third pool was made for primer pairs that commonly had lower depth compared to the average (details Supplementary Table 1). PCR cycling conditions were adapted to the new sequencing primers, with annealing temperature changed to 60°C. Sequencing was carried out with the Oxford Nanopore Technologies MinION sequencer. Samples that had poor coverage (<70%) with the ARTIC primer set were repeated with the UNZA primer set.

### Analysis of SARS-CoV-2 sequencing data

Raw FAST5 data produced by the MinION were processed with Guppy v5.0.7. FAST5s were basecalled with guppy_basecaller, basecalled FASTQs were assigned to barcodes using guppy_barcoder, including the ‘--require_barcodes_both_ends’ flag. The per-sample FASTQ files were processed with the artic pipeline using the ‘medaka’ option [21]. The lineage of each consensus genome was identified using pangolin with the following versions; pangolin v3.1.17, pangolearn 2021-12-06, constellations v0.1.1, scorpio v0.3.16, pango-designation used by pangoLEARN/Usher v1.2.105, pango-designation aliases v1.2.122 [22]. Samples were re-analysed when the Pangolin database was updated. The run was repeated if there was contamination in the negative control.

To set reasonable Ct thresholds for selecting samples to sequence in future work, we plotted the true positive rate versus the false positive rate (i.e. ROC curves) for a range of Ct thresholds from 15 to 40, where the true positive rate was defined as the proportion of samples with a genome coverage >=70% that had a Ct below the threshold. The false-positive rate was defined as the proportion of samples with a genome coverage <70% that had a Ct below the threshold. Code to calculate the values for the ROC curves is available here - https://gist.github.com/flashton2003/bb690261106dc98bb1ae5de8a0e61199. The lineage/VOC of samples in GISAID was obtained via the GISAID website (https://www.epicov.org/epi3/start).

### Statistical analysis

Clinical data were analysed using Stata V15.1 (StataCorp, Stata Statistical Software: Release 15, College Station, Texas, USA). Categorical variables were compared using Fisher’s exact test. Continuous variables were tested for normality and appropriate statistical tests were applied; non-normally distributed measurements are expressed as the median [IQR] and were analysed by the Kruskal-Wallis test to compare clinical parameters across the four waves. The primary outcome variable was survival to hospital discharge. We selected the following covariates **a priori** to determine potential predictors of mortality: pandemic infection wave; vaccine status; age; sex; HIV infection status; prior diagnosis of cardiac disease; prior diagnosis of diabetes mellitus; time from symptoms to hospital admission; respiratory rate; and oxygen saturation (SpO_2_). This information was obtained from the patients admission files, health passport, medical chart or other documents. HIV was not independently confirmed, but was determined from patient medical records. All the above variables were included within the multivariable model and were collected at, or shortly after, hospital admission (selected as clinically relevant parameters that could reasonably be used by clinicians to influence treatment decisions). Univariable and multivariable logistic regression analyses were fitted using the STATA “logistic” command to generate odds ratios and confidence intervals (see supplementary materials). In addition, we conducted an exploratory sensitivity analysis, excluding patients who did require supplemental oxygen (indicative of less severe disease) at the time of enrolment. The overall statistical significance of the difference in mortality between waves was assessed using a likelihood ratio test, comparing the univariable model against a null, intercept-only model and the full multivariable model against a null model with all covariates except for the categorical variable encoding the epidemic wave. Statistical analysis and plotting of genomic results was done using R v4.1.0 [23]. Exact binomial confidence intervals for the proportion of each genotype during each wave were calculated using the binom.test function. Statistical analysis STATA code is available here https://gist.github.com/flashton2003/c241f1153a6a9cb76a26f5857fe53976).

## Results

### Patient Recruitment and SARS-CoV-2 genomic analysis

Between July 2020 and March 2022, we recruited 314 adults with PCR confirmed COVID-19 disease, using the ISARIC Clinical Characterisation Protocol (Table 1). Recruitment spanned four distinct waves of COVID-19 in Malawi; 1^st^ wave n=48 (July-November 2020), 2^nd^ wave n=94 (December 2020-March 2021), 3^rd^ wave n=97 (June 2021-October 2021) and 4^th^ wave n=75 (December 2021-March 2022). The higher number of participants recruited in waves 2 and 3 reflected the epidemiology of COVID-19 in Malawi (Supplementary Figure 1). Overall, 89.5% of patients survived to hospital discharge (per wave numbers can be seen in Table 1).

**Table 1:** Comparison of the demographic and clinical characteristics of COVID patients enrolled in ISARIC during three waves. UVA: Universal Vital Assessment score (16) LOS: length of stay. TB positivity was defined according to presence of positive urinary LAM, GeneXpert or sputum test during hospital admission. Diabetes and Cardiac disease status ascertained from patient history and medical notes. # Proportion (%) positivity calculated using the denominator for individual variables (unknown status classified as missing data) and compared using the Fisher’s exact test. §: Median and IQR were compared using the Kruskal-Wallis test

|  | **W1 - "B1"**<br>**(n=48)** | **W2 - Beta**<br>**(n=94)** | **W3 - Delta**<br>**(n=97)** | **W4 - Omicron**<br>**(n=75)** | **P value** |
| --- | --- | --- | --- | --- | --- |
| Female<sup>§</sup> | 31.3% (15) | 41.5% (39) | 28.9% (28) | 36.0% (27) | 0.302 |
| Age<sup>§</sup> | 52 (43 – 64) | 46 (37 – 58) | 50 (38 – 63) | 42 (34 – 58) | 0.132 |
| Days from symptoms to admission<sup>§</sup> | 5 (2 – 8) | 4 (2 – 9) | 2 (1 – 5) | 2 (0 – 4) | <0.001 |
| Days from admission to sample<sup>§</sup> | 4 (2 – 5) | 3 (2 – 7) | 3 (2 – 5) | 3 (2 – 5) | 0.725 |
| HIV positive | 22.9% (11) | 29.8% (28) | 26.8% (26) | 36.0% (27) | 0.422 |
| TB positive | 2.1% (1) | 1.1% (1) | 1.0% (1) | 1.3% (1) | 1.000 |
| Malaria positive | 4.2% (2) | 2.1% (2) | 1.0% (1) | 0.0% (0) | 0.274 |
| Cardiac disease | 30.0% (13) | 23.4% (22) | 4.1% (4) | 5.3% (4) | <0.001 |
| Diabetes | 40.0% (18) | 19.2% (18) | 17.5% (17) | 6.7% (5) | <0.001 |
| Oxygen on enrolment | 50.0% (23) | 58.5% (55) | 63.9% (62) | 22.7% (17) | <0.001 |
| UVA score<sup>§</sup> | 2 (0 – 4) | 2 (0 – 3) | 2 (0 – 4) | 0 (0 – 2) | 0.001 |
| Beta-lactam antibiotic | 81.3% (39) | 68.1% (64) | 82.5% (80) | 73.3% (55) | 0.096 |
| Steroids | 60.4% (29) | 59.6% (56) | 84.5% (82) | 38.7% (29) | <0.001 |
| Survival to discharge | 91.7% (44) | 90.4% (85) | 83.5% (81) | 94.7% (71) | 0.118 |
| Survivor LOS<sup>§</sup> | 8 (6 – 18) | 8 (4 – 16) | 8 (6 – 11) | 7 (4 – 13) | 0.368 |
| ≥1 Vaccine | 0% (0) | 0% (0) | 21.7% (21) | 20.0% (15) | <0.001 |

The sequencing laboratory received viral material from 161 of 314 participants. RT-PCR Ct values were available for 156 cases. There was no difference between Ct values from the different waves (Supplementary Figure 2, Kruskal-Wallis test P-value 0.24). There was no significant difference between Ct values from patients who were HIV positive, HIV negative, or whose HIV status was unknown (Supplementary Figure 3, Kruskal-Wallis test P-value = 0.22), although measures of the degree of immunosuppression were unavailable.

We sequenced all samples with a Ct below 27 (this cut-off was selected based on Supplementary Figure 4), and as many samples with a Ct above 27 as sequencing capacity allowed. Of the 161 cases for which we received viral material, we sequenced 126 samples from 126 patients and obtained 55 genomes with greater than 70% coverage at 20x depth (Supplementary Table 2). Low coverage of the genome (<70%) was associated with low viral load (i.e., high Ct). This was true for both ARTIC v3 and UNZA tiling PCR primer sets (Figure 1). Overall, the median Ct value of samples with <70% coverage at 20x depth was 32.0, compared with a Ct 25.9 for samples with >=70% coverage (Supplementary Tables 2 & 3).

**Figure 1:**
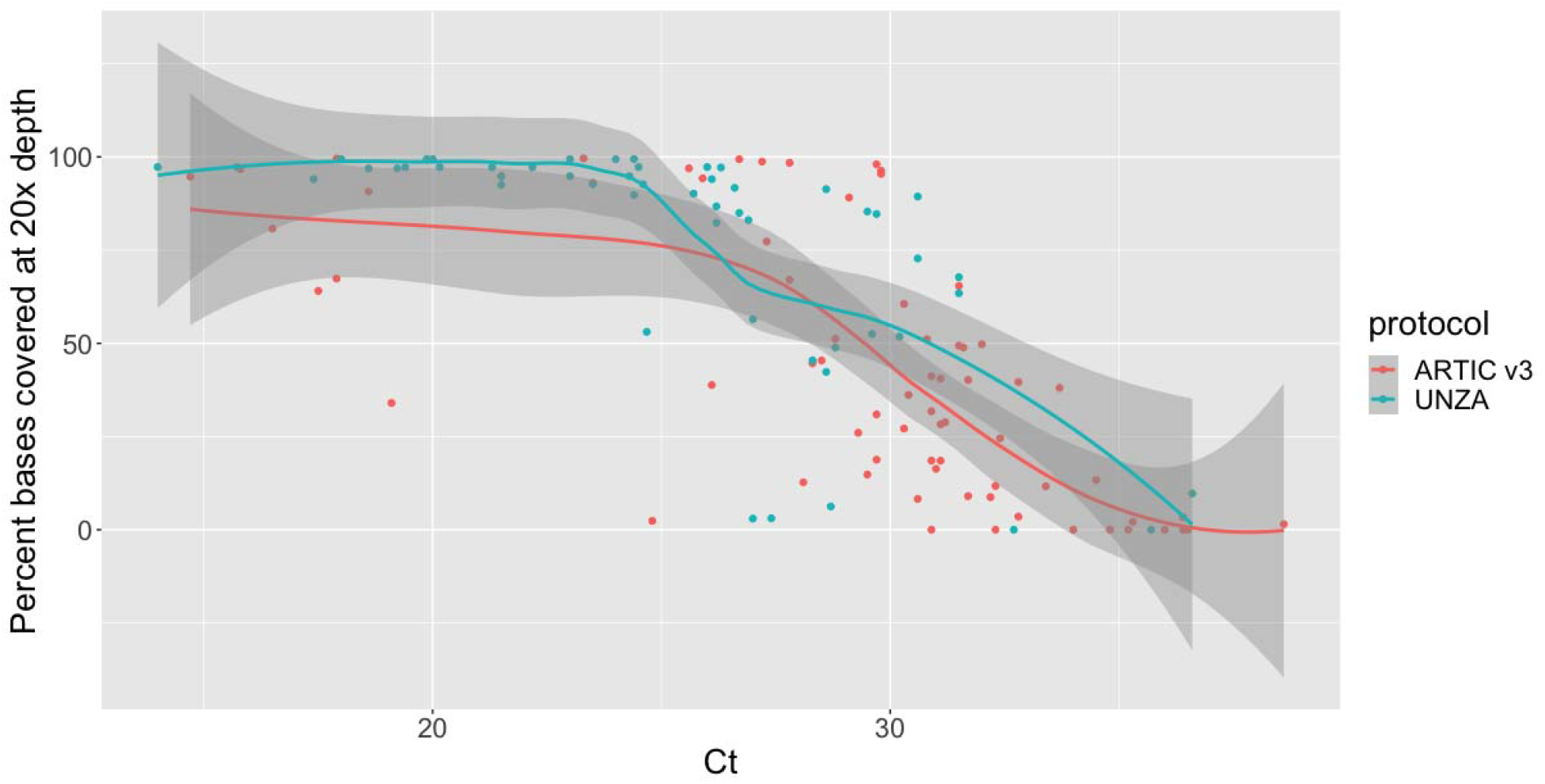
Relationship between PCR Ct value and the percentage of the SARS-CoV-2 reference genome covered to at least 20x depth. The number at the top of each column is the number of samples for the two protocols in each bin of the box plot.

We observed three lineages among the 11 SARS-CoV-2 samples from wave 1 (Figure 2, Supplementary Table 2), with the most frequently identified pangolin lineage being B.1 (n=8), followed by B.1.1 (n=2) and B.1.1.448 (n=1). All 6 samples from wave 2 were VOC Beta (exact binomial 95% CI of the estimate in the untested population = 54-100%) and 96% (25/26) of samples from wave 3 were VOC Delta (95% CI 80-100%) (Figure 2). One sample received at the beginning of June 2021 was VOC Beta. We observed seven pangolin lineages among the 25 VOC Delta samples sequenced during wave 3; AY.75.1 (n=11), B.1.617.2 (n=8), AY.75 (n=2) and 1 each of AY.50, AY.59, AY.122 and AY.72 (Supplementary Figure 5). Of the 12 successfully sequenced samples from wave 4, 100% (95% CI 73.5-100%) were Omicron VOC. Eleven of twelve were BA.1 with the remaining sample belonging to BA.2. The BA.2 sample came from a patient enrolled in February 2022. Due to low numbers of successfully sequenced isolates during the second wave, we also obtained the genotype of samples from Malawi submitted to GISAID during this time, for which explicit permission could be obtained for re-use from the data depositor; Beta VOC accounted for 100 of the 104 (96%, 95% CI: 90-98%) SARS-CoV-2 genomes from Malawi in GISAID which were sampled.

**Figure 2:**
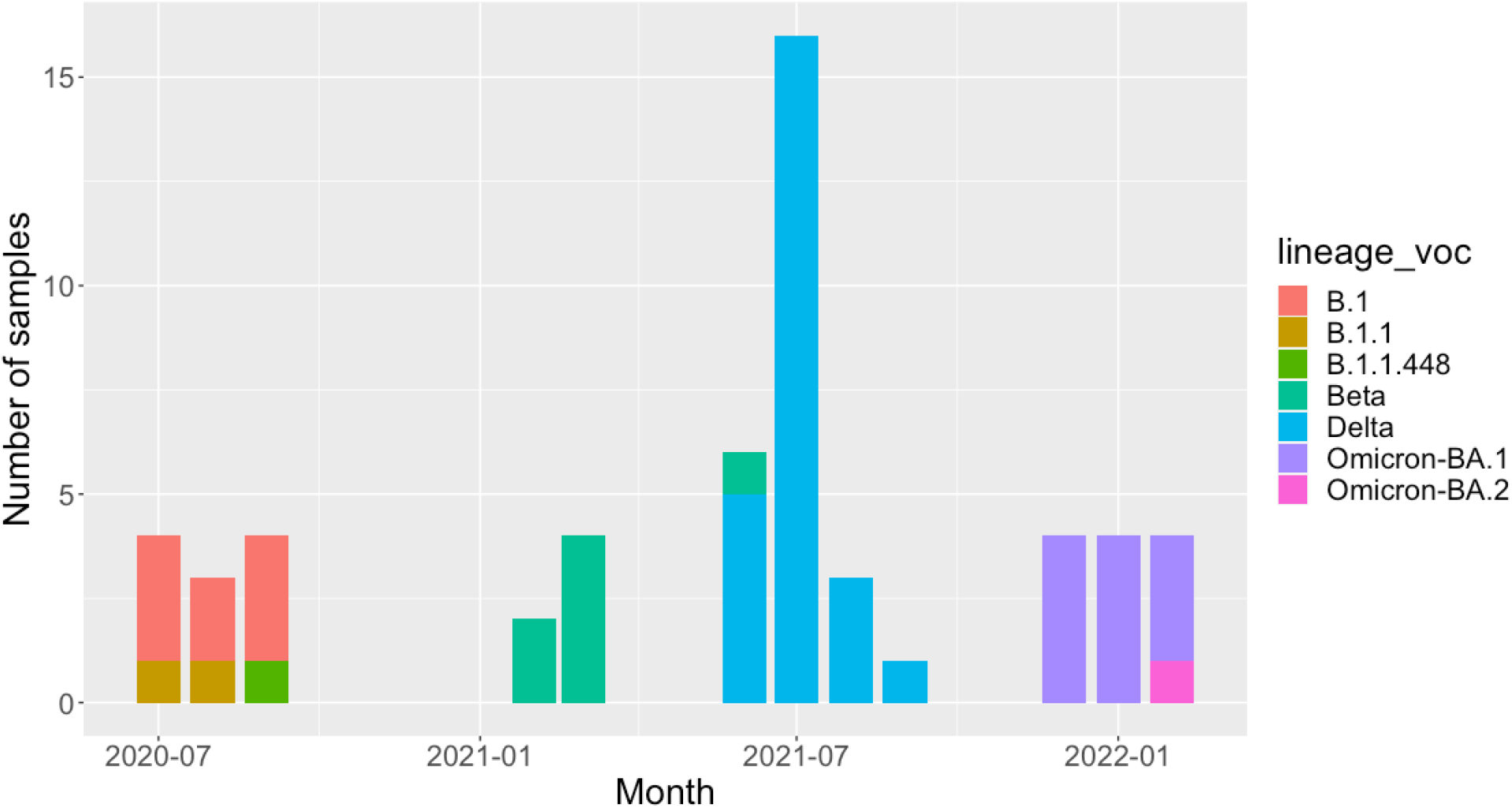
The monthly number of each lineage or VOC identified in patients in our cohort.

### Clinical Characteristics

There were no significant differences in sex or median age between participants between waves (Table 1), however, there was a significant reduction (p=<0.001) in time from symptom onset to presentation in Delta (median two days [IQR 1-5]) and Omicron waves (median two days [IQR: 0-4]) compared to the B.1 (median five days [IQR: 2-8]) or Beta waves (median four days [IQR: 2-9]). There was a lower percentage of patients with cardiac disease (30.0% and 23.4% vs 4.1% vs 5.3%, P <0.001) and diabetes (40% vs 19.2% vs 17.5% vs 6.7% p=<0.001) in later waves. There was a significant reduction in the numbers of patients requiring oxygen at enrolment during the Omicron wave, with the highest proportion during Delta wave (50% vs 58.5% vs 63.9% vs 22.7% p=<0.001). Similarly, fewer patients were given steroids during Omicron wave, with the highest numbers receiving steroids in Delta wave (60.4% vs 59.6% vs 84.5% vs 38.7% p=<0.001). Overall, few patients were vaccinated; in this cohort 21/97 (21.7%) Delta wave participants and 15/75 (20%) Omicron wave participants had received at least one dose of any vaccine. For both unvaccinated and vaccinated groups survival was just under 90% (p=0.9).

Univariable logistic regression analysis demonstrated that age ≥70 (OR7.21 CI:1.48-35.07), respiratory rate ≥ 30 (OR 14.87 CI: 3.09 – 71.71) and SpO_2_ ≤87% (OR 15.4 CI: 5.66– 41.93) were associated with mortality, although with wide confidence intervals (Table 2). Multivariable analysis showed a statistically significant increase in case fatality rate in the whole cohort during the Delta wave (OR 4.99 CI 1.00-25.02) (Table 2). However, the likelihood ratio test for the presence or absence of wave within the model was not significant (Chi2 = 5.91, p = 0.116).

**Table 2:** Clinical factors associated with mortality for SARS-CoV-2 PCR confirmed patients admitted to hospital with severe acute respiratory infection. Univariable and multivariable logistic regression analysis with all pre-specified parameters included within the final multivariable model. Final multivariable model: n=226, chi^2^ = 62.80, Pseudo R^2^ = 0.363.

| Variable | Univariate |  |  | Multivariate |  |  |
| --- | --- | --- | --- | --- | --- | --- |
|  | Odds ratio | P value | Confidence Interval | Odds ratio | P value | Confidence Interval |
| Wave |  |  |  |  |  |  |
| 2 | 1.16 | 0.808 | 0.34 – 4.00 | 1.38 | 0.686 | 0.29 – 6.51 |
| 3 | 2.17 | 0.188 | 0.68 – 6.90 | 4.99 | 0.050 | 1.00 – 25.02 |
| 4 | 0.62 | 0.514 | 0.15 – 2.61 | 2.24 | 0.392 | 0.35 – 14.16 |
| Vaccinated | 1.07 | 0.900 | 0.35 – 3.25 | 0.92 | 0.916 | 0.21 – 4.10 |
| Age |  |  |  |  |  |  |
| 30-39 | 0.66 | 0.679 | 0.09 – 4.85 | 0.25 | 0.262 | 0.02 – 2.83 |
| 40-49 | 3.22 | 0.145 | 0.67 – 15.51 | 1.54 | 0.627 | 0.27 – 8.86 |
| 50-59 | 1.38 | 0.717 | 0.24 – 7.93 | 0.51 | 0.559 | 0.05 – 4.85 |
| 60-69 | 1.90 | 0.473 | 0.33 – 10.98 | 0.76 | 0.795 | 0.09 – 6.31 |
| ≥70 | 7.21 | 0.014 | 1.48 – 35.07 | 9.55 | 0.026 | 1.31 – 69.77 |
| Male | 0.60 | 0.174 | 0.29 – 1.25 | 0.51 | 0.190 | 0.19 – 1.39 |
| HIV positive | 0.82 | 0.654 | 0.33 – 1.99 | 1.08 | 0.898 | 0.32 – 3.65 |
| HIV unknown | 1.28 | 0.573 | 0.54 – 3.07 | 0.96 | 0.946 | 0.30 – 3.11 |
| Cardiac disease | 1.44 | 0.456 | 0.56 – 3.71 | 0.82 | 0.792 | 0.19 – 3.51 |
| Diabetes | 1.20 | 0.690 | 0.49 – 2.91 | 1.15 | 0.818 | 0.35 – 3.83 |
| Symptoms to admission (days) |  |  |  |  |  |  |
| 4-6 | 2.64 | 0.037 | 1.06 – 6.58 | 2.56 | 0.132 | 0.75 – 8.67 |
| 7-9 | 2.59 | 0.101 | 0.84 – 8.06 | 4.24 | 0.098 | 0.77 – 23.49 |
| ≥10 | 2.19 | 0.127 | 0.80 – 5.99 | 2.70 | 0.160 | 0.68 – 10.75 |
| Respiratory rate |  |  |  |  |  |  |
| 20-24 | 2.18 | 0.321 | 0.47 – 10.13 | 1.28 | 0.778 | 0.23 – 7.10 |
| 25-29 | 4.07 | 0.084 | 0.83 – 20.02 | 1.16 | 0.874 | 1.78 – 7.62 |
| ≥30 | 14.87 | 0.001 | 3.09 – 71.71 | 5.97 | 0.067 | 0.88 – 40.26 |
| SpO2 |  |  |  |  |  |  |
| 93-95 | 1.39 | 0.569 | 0.45 – 4.30 | 0.74 | 0.659 | 0.20 – 2.80 |
| 88-92 | 2.54 | 0.093 | 0.86 – 7.53 | 1.44 | 0.569 | 0.41 – 5.01 |
| ≤87 | 15.40 | <0.001 | 5.66 – 41.93 | 11.22 | 0.001 | 2.59 – 48.65 |

Therefore, these exploratory findings within our limited cohort should not be overinterpreted. HIV infection; presence of co-morbidities; days from symptoms to admission; and respiratory rate were not associated with survival within the multivariable model. We conducted an exploratory sensitivity analysis including only participants who required oxygen at study enrolment as a marker of disease severity (n=157, of whom 26 [16.6%] died). This demonstrated that admission during Delta wave was independently associated with mortality within a multivariable analysis (OR 13.91 [CI: 1.56-125.06, p=0.018) (Supplementary Table 4).

## Discussion

Using genomic sequencing we were able to define the viral sub-types or VOCs associated with four distinct waves of patients hospitalised with COVID-19. The first wave was predominantly B.1, all sequenced samples from the second wave were Beta VOC, the sequenced samples from the third wave were predominantly Delta, whilst the samples from the fourth wave were largely Omicron BA.1. Infection with Delta variant was associated with a higher risk of mortality, particularly in patients requiring oxygen during admission. This study reports clinical differences in outcome between SARS-CoV-2 variants in a low-income southern African setting in a population with a high burden of infectious disease, including HIV.

The increased risk of mortality in this cohort was associated with increased age (≥70 years) and low oxygen at recruitment (SpO2 <87%), in line with other cohorts (ISARIC, [24]). While our small sample size necessitates caution in interpretation, there was an increased risk of death associated with Delta VOC, particularly in those patients requiring oxygen. Increased mortality with Delta VOC has been reported elsewhere [13–16], but not consistently in Africa [25], where robust clinical data has not commonly been linked with SARS-CoV-2 sequencing data. Patients with severe disease were managed with oxygen, steroids and beta-lactam antibiotics, consistently applied within the hospital between waves. We did not observe an excess of deaths in people living with HIV, however the sample size was low and we did not assess level of immune-suppression in these patients [26]. Patients admitted during the Omicron wave required less oxygen at enrolment, suggesting they were less unwell at presentation, although overall mortality was not significantly lower. This is consistent with other studies in sub-Saharan Africa where patients admitted with COVID-19 during Omicron waves had comparatively less severe disease [16,27,28]. There is a high burden of HIV and a low SARS-CoV-2 vaccine coverage in Malawi [29], this provides a plausible environment for the emergence of novel VOCs [30–33]. It is crucial to identify potential VOCs rapidly and report these internationally. The continuation of in-country genomic surveillance in Malawi is therefore important locally and globally.

Throughout the study there was no invasive and very limited non-invasive ventilatory support available for COVID-19 patients and no access to newer therapies such as interleukin-6 antagonists. Therapeutic options for COVID-19 in high income settings are developing rapidly, with genomic viral sequencing used to guide treatments (<u>NICE</u>). This study thus highlights significant inequity in availability of globaly recommended therapeutics for COVID-19 despite relatively high rates of in-patient mortality. It is unclear from this study whether the reduction in severity seen in the Omicron wave was affected by immunity – either vaccine derived or naturally acquired. Overall, 20.9% of the recruited patients in waves three and four were vaccinated with at least one dose (predominantly Astra-Zeneca ChAdOx1-S and J&J Ad26.COV2.S), which is higher than the background population overall, but similar to rates seen in urban Blantyre (25% at least one dose by Feb 2022, Personal Communication, Blantyre District Health Office). However there were already high rates of sero-positivity amongst blood donors in Malawi with 70% of adults SARS-CoV-2 sero-positive in July 2021 during the Delta wave [34] suggesting high population exposure with naturally acquired immunity.

A strength of our study is that we carried out sequencing and analysis in Malawi directly linked with robust and systematically collected clinical data. In country analysis allowed us to report our findings to clinical and public health partners rapidly. Vital to our success in establishing sequencing in Malawi was the portability of the MinION sequencer; the public lab protocols (18); bioinformatics software from the scientific community (13); and the infrastructure and funding available to us as an international research institution. The MinION platform has become intergral to outbreak response, as demonstrated for SARS-CoV-2 (19,20), Ebola (21) and Zika (22). However, even with this portable and low-maintenance sequencer (with no service contracts or engineer visits required); experienced molecular biologists and bioinformaticians; and considerable international support, it was still very difficult to establish sequencing capability. In particular, we found it extremely challenging to procure reagents, and this was exacerbated by border closures and travel restrictions. As there is no existing policy framework within Malawi for the integration of sequencing data into public health decision making, the utility of our data to decision makers was limited.

Our study has several limitations. We produced a relatively small number of sequences. This was partly due to the limited number of patients recruited into the study during each wave but also because patients frequently presented with Ct values too high to generate good quality sequence data. Secondly, our observations are limited to a sample of hospitalised patients in a single centre in the southern region of Malawi. Our relatively low sample size impairs our ability to draw firm conclusions on the association between wave and patient outcome. Finally, we recognize that we may not be capturing the full diversity of SARS-CoV-2 circulating in the community, as our sampling of hospitalised patients represents a considerable bias towards people with severe disease, and there is likely to be significant under ascertainment of cases [34].

In conclusion, pragmatic clinical research protocols coupled with portable sequencing capacity enabled us to improve our understanding of the clinical characteristics and impact of the multiple waves of COVID-19 pandemic in Malawi. We recommend that funders support the development of capacity in genomic surveillance of agents of communicable disease, focussing their strategies on endemic diseases, which can pivot to pandemics and outbreak scenarios as the need arises. A key part of this is the development of robust networks for the production and distribution of molecular biology reagents, mirroring what is being developed for vaccines, as this would enable a more rapid and sustained response to future pandemics. Challenges and opportunities arising from this work are detailed in Box 1. Data and sample collection was enabled by collaboration with the ISARIC consortium. This enabled us to enrol patients very quickly using tools already developed for pandemic response. We were also able to contribute valuable clinical data from a low income setting to global analyses.

## Data Availability

All genome sequences are available in GISAID. Accessions are available in Supplementary Table 2.

## Acknowledgments

The authors thank all study participants and the staff of the Queen Elizabeth Central Hospital (QECH) for their support and co-operation during the study. We would like to thank all the people mentioned in Supplementary File 1 for sharing their data to GISAID.

This work was supported by the UK Foreign, Commonwealth and Development Office and Wellcome grants for SARS-CoV-2 diagnostics [220757/Z/20/Z] and the MLW Core grant [206545/Z/17/Z]. KGB is supported by an NIH-Fogarty fellowship [TW010853]. The study was reported in line with STROBE guidelines.

We would like to acknowledge the contribution of the following members of the Blantyre COVID-19 Consortium; Clinical - Wezzie Kalua, Peter Mandala, Barbara Katutula, Rosaleen Ng’oma, Steven Lanken, Jacob Phulusa, Mercy Mkandawire, Sylvester Kaimba, Sharon Nthala, Edna Nsomba, Lucy Keyala, Beatrice Chinoko, Markus Gmeiner, Vella Kaudzu, Bridget Freyne, Todd D. Swarthout and Pui-Ying Iroh Tam. Laboratory - Simon Sichone, Ajisa Ahmadu, Grace Stima, Mazuba Masina, Oscar Kanjewa, Vita Nyasulu, End Chinyama, Allan Zuza, Brigitte Denis, Evance Storey, Nedson Bondera, Danford Matchado, Adams Chande, Arthur Chingota, Chimenya Ntwea, Langford Mkandawire, Chimwemwe Mhango, Agness Lakudzala, Mphatso Chaponda, Percy Mwenechanya, Leonard Mvaya and Dumizulu Tembo. Data and statistics - Marc Y. R. Henrion, James Chirombo, Paul Kambiya, Clemens Masesa & Joel Gondwe.

## Data availability statement

All genome sequences are available in GISAID and INSDC databases – accessions are available in Supplementary Table 2.

