## Supplementary Figures for "A comparison of four epidemic waves of COVID-19 in Malawi; an observational cohort study"

### Slide 1
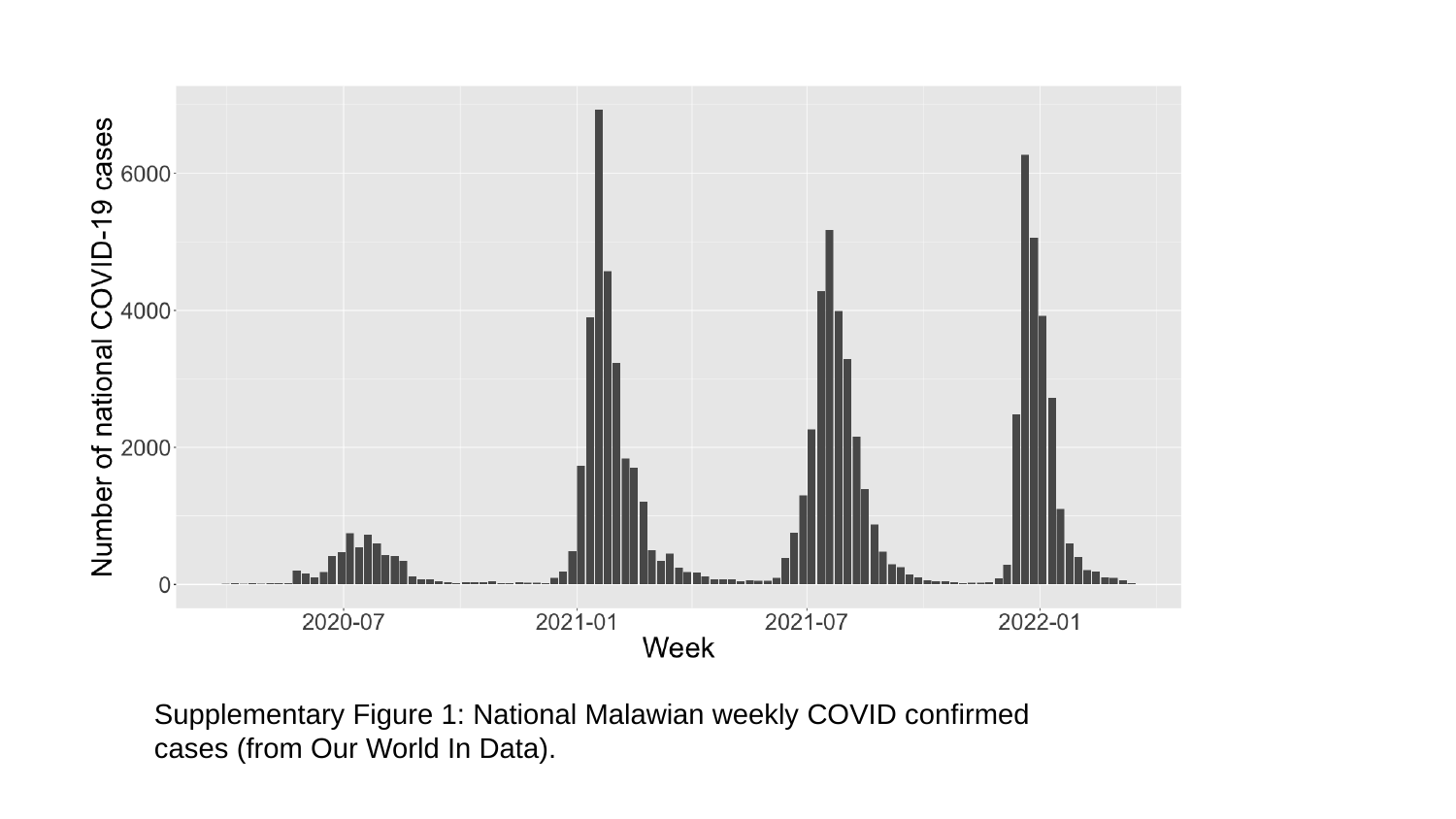

Supplementary Figure 1: National Malawian weekly COVID confirmed cases (from Our World In Data).

### Slide 2
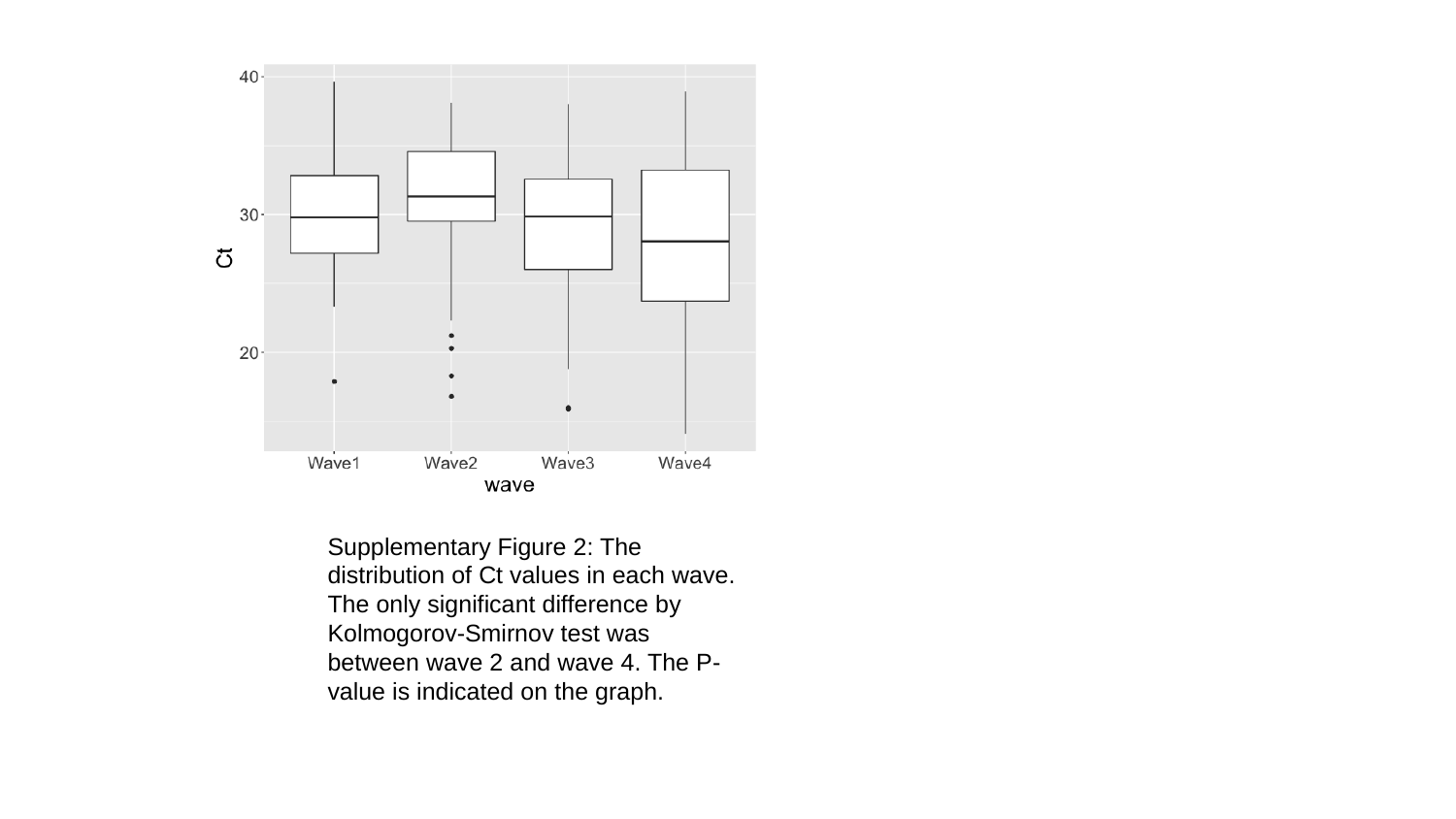

Supplementary Figure 2: The distribution of Ct values in each wave. The only significant difference by Kolmogorov-Smirnov test was between wave 2 and wave 4. The P-value is indicated on the graph.

### Slide 3
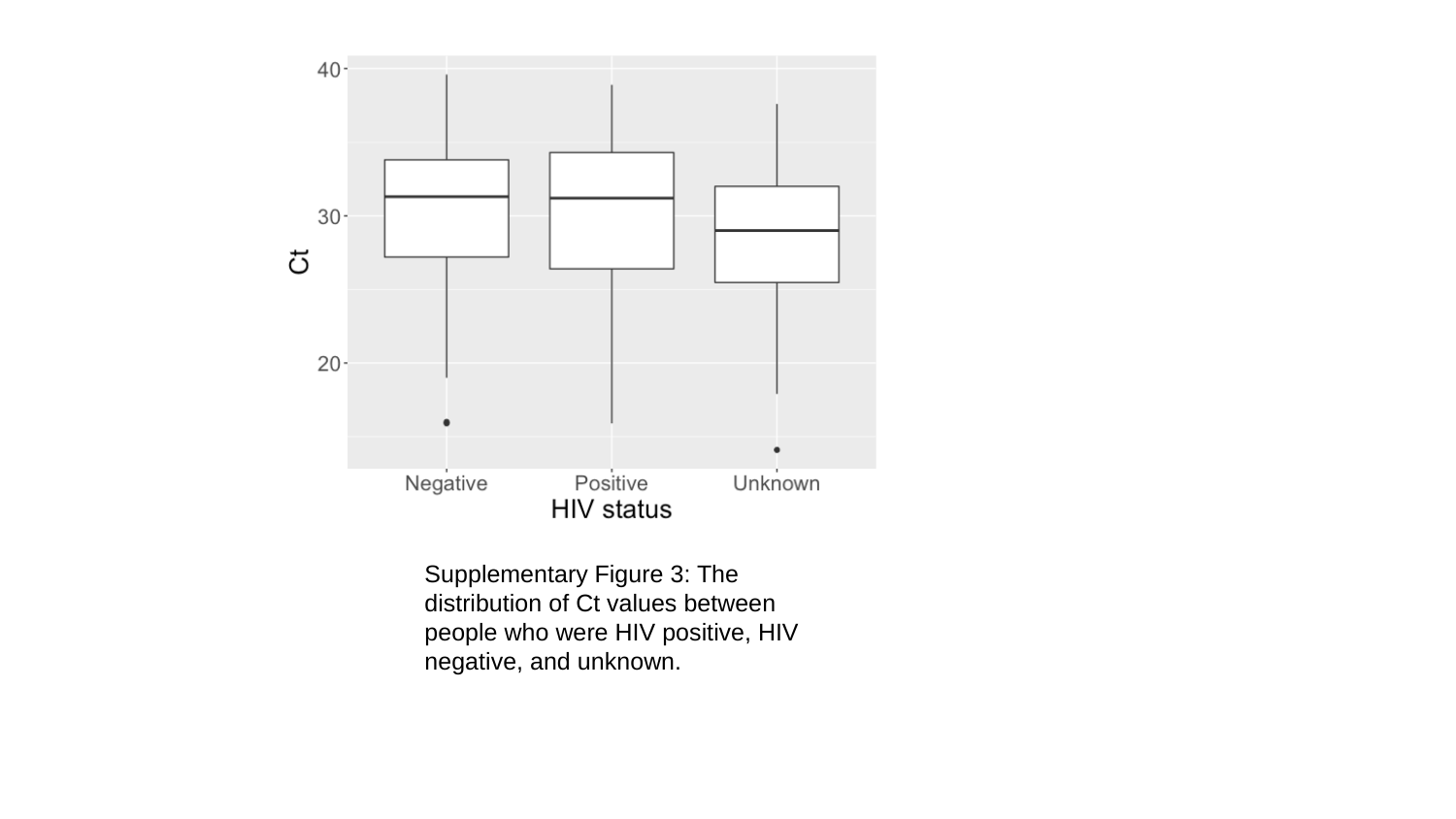

Supplementary Figure 3: The distribution of Ct values between people who were HIV positive, HIV negative, and unknown.

### Slide 4
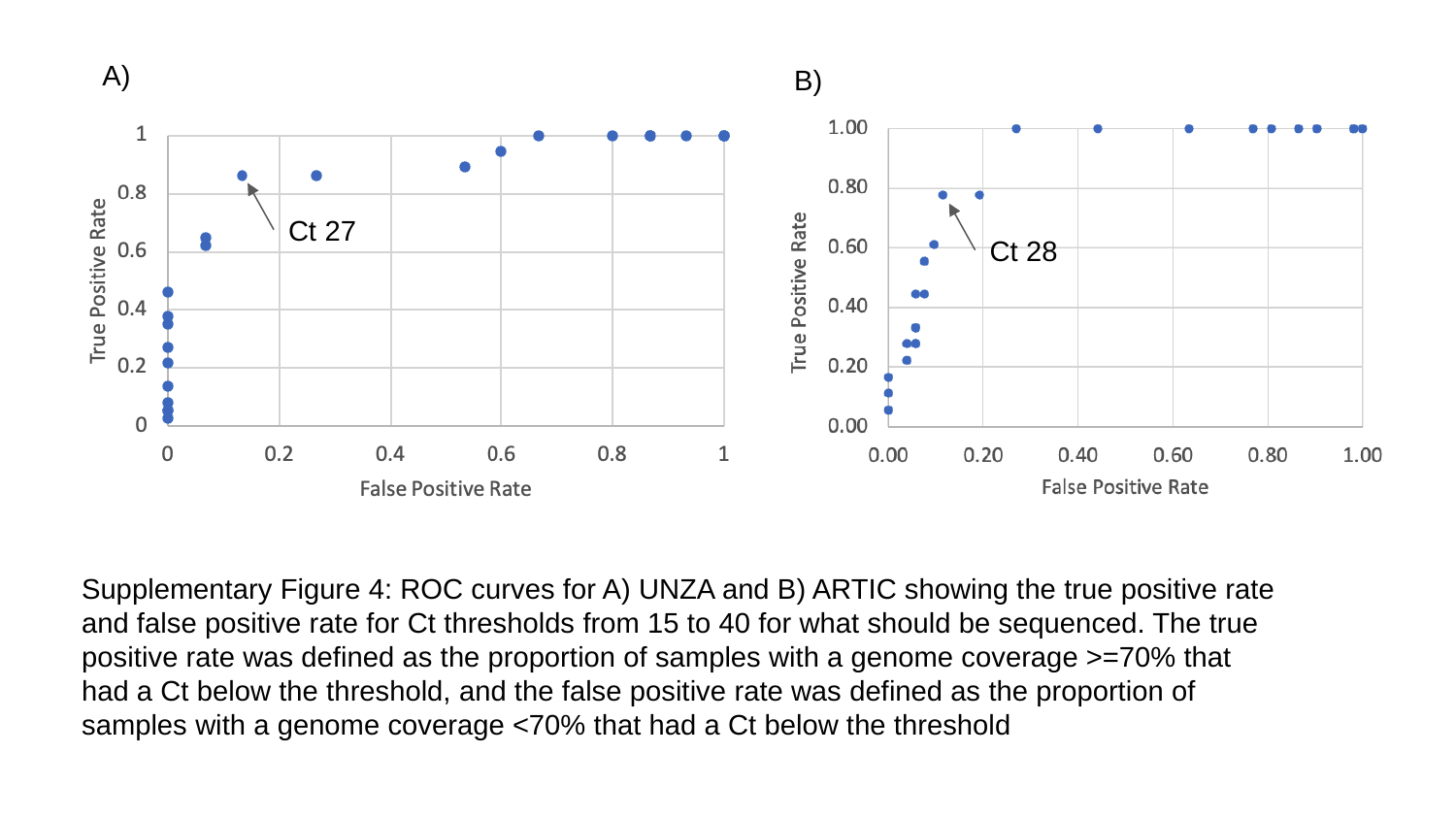

A)
B)
Ct 27
Ct 28
Supplementary Figure 4: ROC curves for A) UNZA and B) ARTIC showing the true positive rate and false positive rate for Ct thresholds from 15 to 40 for what should be sequenced. The true positive rate was defined as the proportion of samples with a genome coverage >=70% that had a Ct below the threshold, and the false positive rate was defined as the proportion of samples with a genome coverage <70% that had a Ct below the threshold

### Slide 5
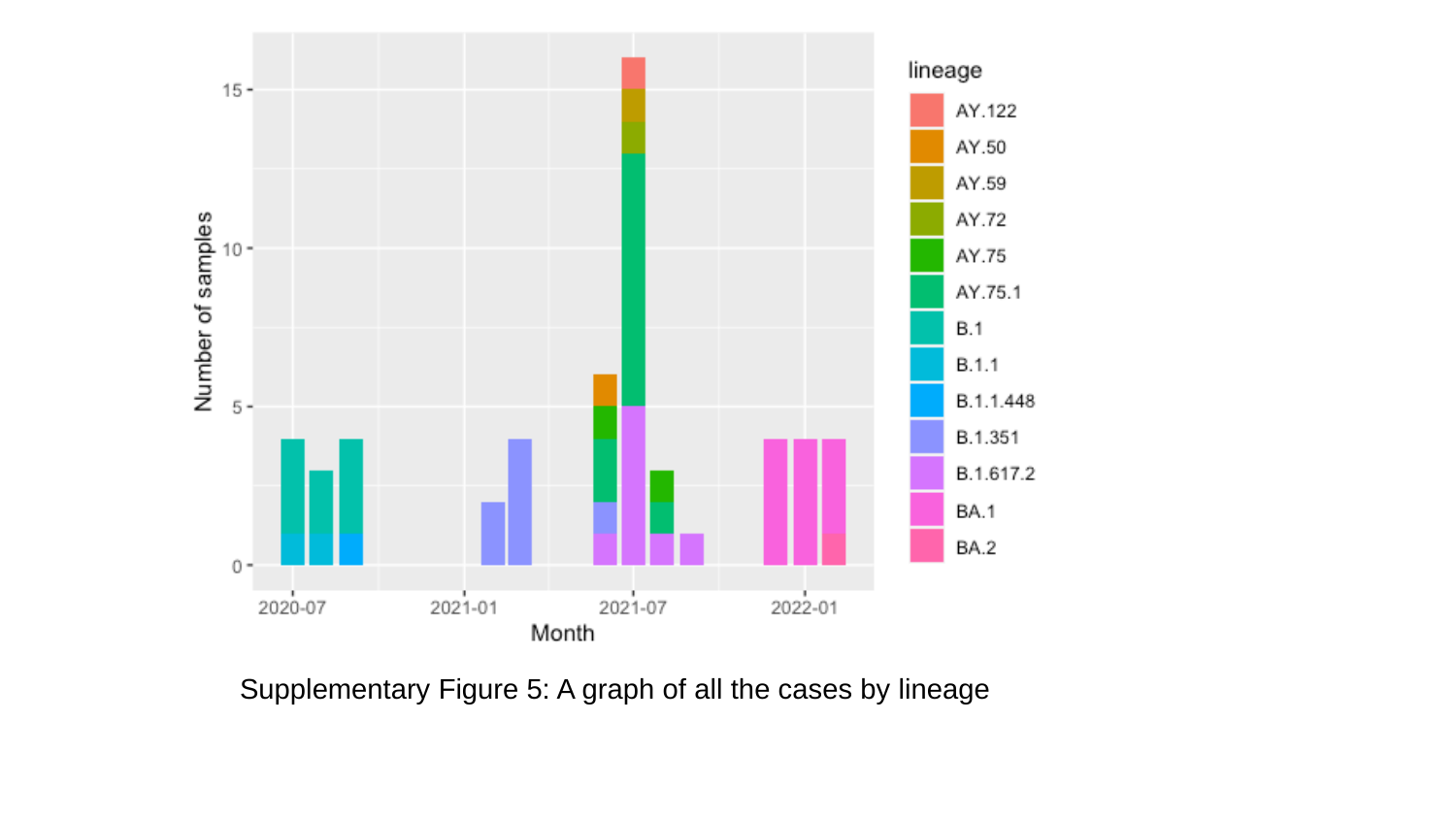

Supplementary Figure 5: A graph of all the cases by lineage
